# Coffee, smoking and aspirin are associated with age at onset and clinical severity in idiopathic Parkinson’s disease

**DOI:** 10.1101/2021.11.08.21265971

**Authors:** Carolin Gabbert, Inke R. König, Theresa Lüth, Beke Kolms, Meike Kasten, Eva-Juliane Vollstedt, Alexander Balck, Fox Insight Study, Anne Grünewald, Christine Klein, Joanne Trinh

## Abstract

Parkinson’s disease (PD) is a progressive neurodegenerative disorder. Genetic modifiers, environmental factors and gene-environment interactions have been found to modify PD risk and disease progression. The objective of this study was to evaluate the association of smoking, caffeine and anti-inflammatory drugs with age at onset (AAO) and clinical severity in a large PD cohort. A total of 35,963 American patients with idiopathic PD (iPD) from the Fox Insight Study responded to health and lifestyle questionnaires. We compared the median AAO between different groups using the non-parametric Mann-Whitney U test. Non-parametric Spearman correlation was used for correlation assessments and regression analysis was used to assess interaction between variables. Reported p-values remain descriptive because they are not corrected for multiple testing and results are exploratory. We found that smoking (r=0.08, p<0.0001), coffee drinking (r=0.69, p<0.0001) and aspirin intake (r=0.23, p<0.0001) show an exploratory association with AAO in iPD. However, the effect of aspirin diminished as an independent predictor after including comorbidities (heart diseases and arthritis). Smoking was associated with higher (more severe) motor scores, while coffee drinking was linked to lower (less severe) motor scores (p<0.05). In addition, smokers reported anxiety, depression and other non-motor symptoms such as unexplained pains and problems remembering (p<0.05). The association of aspirin with PD AAO was replicated in another cohort (EPIPARK) (n=237 patients with PD), although again the effect diminished after including age in the regression model. Future longitudinal studies are warranted to investigate the clinical severity over time.

## Introduction

Parkinson’s disease (PD) is a progressive neurodegenerative disorder, characterized by dopaminergic neuronal loss in the substantia nigra and the presence of Lewy Bodies [1, 2]. It is the second most common neurodegenerative disorder and the fastest growing neurological disease currently affecting over 7 million patients worldwide [3].

A phenomenon in PD is variable age at onset (AAO) that is considered a consequence of genetic and environmental factors. Tobacco use and smoking are already known protective factors for PD risk [4-6]. However, research specifically on AAO is not as extensive. Studies report that disease onset in patients with idiopathic or monogenic PD is later among smokers, dependent on the dosage [7-13]. The largest cross-sectional cohort comprised 512 PD patients, of whom 184 were smokers and 328 never smoked [10]. Likewise, caffeine consumption was associated with lower PD risk, with a dosage-dependent level of protection [14]. In terms of AAO, there is evidence that the onset of PD among coffee drinkers is later compared to non-drinkers [11, 15-17], also indicating a dosage effect [12, 18]. However, earlier studies report opposing effects of an earlier AAO with higher coffee intake [7]. Non-steroidal anti-inflammatory drug (NSAID) intake has been found to be associated with a lower risk for PD [19], supporting work that describe a role for neuroinflammatory signaling in PD [20]. NSAIDs (ibuprofen and aspirin) have been found to influence the penetrance of LRRK2 [21]. However, there are currently no studies published that investigate an association between aspirin and AAO in iPD. The impact of environmental and lifestyle factors on AAO and disease-related symptoms of PD still have not been investigated in large cohorts. Thus far, 13 studies analyzed the effect of tobacco or caffeine on PD AAO [7, 8, 10-13, 15-18, 22-24] (Fig. S1 and Table S1). These cross-sectional studies have a patient sample size ranging from n=83 to n=512.

Herein, we focused on lifestyle factors implicated in PD risk and investigated the association of smoking, the consumption of caffeine and the use of aspirin on AAO in patients with idiopathic PD (iPD). We hypothesize these factors are associated with AAO in iPD and may be related to motor and non-motor symptoms in a large cohort of American iPD patients (n=35,936).

## Method

### Demographics and participant examination

Our study is composed of 35,963 American patients with PD (Table S2) from the Fox Insight Study (Supplementary text). Most of the patients were White/Caucasian (89.9%) (Table S2). PD patients had a mean age at examination (AAE) of 65.7 ± 10.2 SD years (range: 13.8-119.0 years) and a mean AAO of 60.4 ± 11.0 SD years (range: 5.1-115.4 years); 40.4% of PD patients were female. Patient recruitment for the Fox Insight Study has been previously described [25]. Data from a separate replication cohort of German iPD patients (EPIPARK) was used to test novel associations [26]. In the EPIPARK cohort, PD patients had a mean AAE of 67.7 ± 10.3 SD years (range: 30.0-90.0 years) and a mean AAO of 54.8 ± 13.2 SD years (range: 13.0-81.0 years); 37.3% of PD patients were female. Participant questionnaires (MDS-UPDRS II, NMSQ, GDS, PD-RFQ-Us) are described in detail in the Supplementary text.

### Lifestyle factors

Patients were classified as tobacco users, if they smoked more than 100 cigarettes in their lifetime or if they smoked at least one cigarette per day over a minimal period of six months or if they used smokeless tobacco at least once per day for more than six months. Patients were classified as coffee consumers if they regularly drank caffeinated coffee at least once per week over a period of at least six months. The same classification was used for caffeinated black tea. Lastly, patients were classified as aspirin users if they took at least two pills per week over a minimum of six months.

Duration of smoking, caffeine consumption and aspirin intake were estimated according to the age the patients started using either substance subtracted from the age at termination. If the patients terminated the consumption after their AAO, the age the patients started was subtracted from their AAO. Periods where the patients stopped regularly consuming were not included in the duration. Smoking dosage was estimates as cigarettes smoked per day within smoking duration time excluding implausible values, so that only values lower than 100 cigarettes per day were included in the analyses. Coffee and black tea dosage was defined as cups per week the patients drank within drinking duration time, excluding all values higher than 100 cups per week from the analysis. Aspirin dosage was defined as pills per week the patients took within aspirin intake duration time. The number of cigarettes for non-smokers, cups of coffee or black tea for non-drinkers and pills per week for aspirin non-users was set to zero.

### Statistical analysis

For statistical analyses, non-parametric Mann-Whitney U test was performed to compare the distribution of AAO between different groups. For correlation analyses, non-parametric Spearman correlations and linear regression analyses were used to assess correlations and interactions between variables (GraphPad Software Inc., San Diego, CA, USA). Various multilinear regression models were used to investigate the relationship between environmental factors, age, disease duration and motor/non-motor symptoms (IBM SPSS Statistics, Stanford, CA, USA) (Supplementary text). Reported p-values remain descriptive because they are not corrected for multiple testing and results are exploratory.

## Results

### Smoking

Patients with iPD, who reported use of tobacco, had a later AAO (n=2148; median AAO=63.5 years; IQR=56.1-69.1) compared to non-users (n=3375; median AAO=60.8 years; IQR=53.7-66.7) (p<0.0001) (Fig. 1A and Table S3). Investigation of possible smoking dosage effects on AAO showed that the number of cigarettes per day was associated with later AAO (n=4399, r=0.08, p<0.0001) (Fig. 1B), despite an only small effect size. Similarly, a longer duration of smoking showed a positive correlation with AAO (n=912, r=0.07, p=0.0328) (Fig. 1C), but again with a small effect size.

We investigated whether AAE contributed to the correlation between smoking and AAO. When modeled in a linear regression to predict AAO, both AAE (p<1×10^−5^, β>0.9277, SE<0.0169) and smoking dosage (p=0.0017, β=0.0172, SE=0.0055) remained in the model as independent predictors, but the smoking duration (p=0.5591, β=0.0074, SE=0.0127) did not.

**Fig. 1.**
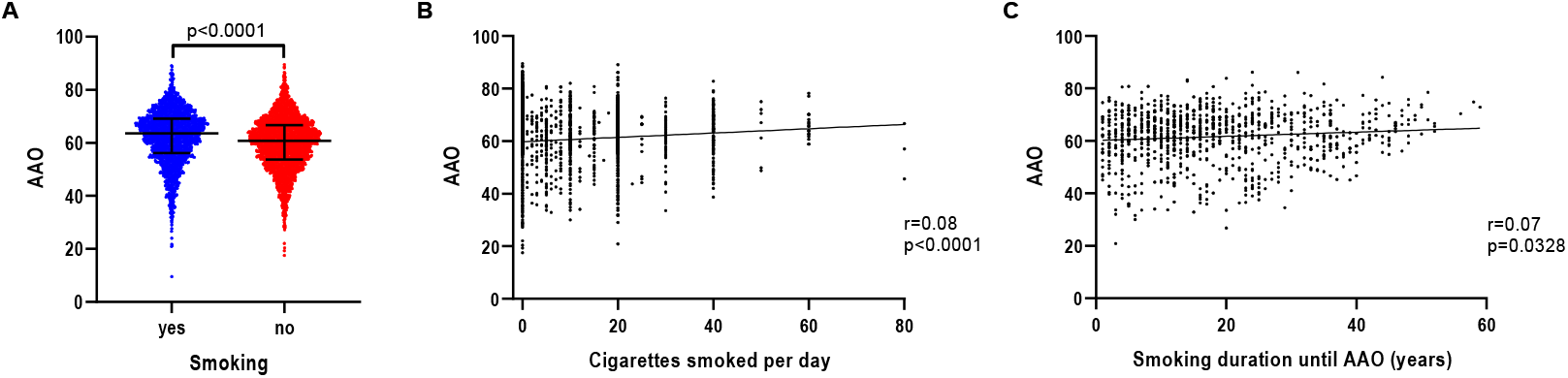
Association of AAO and tobacco use, smoking intensity and smoking duration in iPD. **(a)** Scatter plot of AAO of patients with iPD stratified by smoking status. Median values and interquartile ranges (IQR) are depicted. **(b)** Correlation between number of cigarettes smoked per day and AAO of patients with iPD. **(c)** Correlation between number of years of smoking until AAO and AAO of patients with iPD. P-value: Exploratory Mann Whitney U-test was performed for pairwise comparisons; non-parametric Spearman correlation and simple linear regression analyses were used to assess interactions between variables; p = Spearman’s exploratory p-value, r= Spearman’s rank correlation coefficient

To evaluate more potential predictors of AAO we performed a sensitivity analysis. When modeled in a linear regression to predict AAO (Supplementary text), with covariates smoking duration/dosage, AAE, gender, disease duration (time between AAO and current age) and lung diseases, a negative relationship with smoking duration (p=0.0030, β=-0.0103, SE=0.0034) was observed. However, smoking dosage did not predict AAO (p=0.4208, β=-0.0011, SE=0.0013). A positive relationship was shown for AAO with AAE (p<1×10^−5^, β>0.9900, SE<0.0047) and a negative relationship with disease duration (p<1×10^−5^, β<-0.9519, SE<0.0037) was also present. We also tested for lung diseases including chronic obstructive pulmonary disease (COPD) as potential comorbidity, but these did not show any association with AAO (p>0.1934, β<-0.0262, SE<0.1162).

Given the descriptive associations for AAO, we further investigated the impact of smoking on self-reported motor features (Table 1). When modeled in a logistic regression to predict the occurrence of motor symptoms (Supplementary text and Table S4), with covariates smoking (binary)/duration/dosage, AAE, gender, disease duration (time between AAO and current age) and lung diseases, smoking showed a positive relationship with saliva and drooling (p=0.0165, β=0.0336, SE=0.0140), chewing and swallowing (p=0.0004, β=0.0478, SE=0.0134), and freezing (p=0.0272, β=0.0277, SE=0.0125), when smoking was used as a binary yes-no indication (Table S5). To assess dosage effects, cigarettes smoked per day were used as continuous variable for which non-smokers were set to zero. Smoking dosage demonstrated a positive relationship with chewing and swallowing (p=0.0004, β=0.0018, SE=0.0005), walking and balance (p=0.0320, β=0.0011, SE=0.0005), freezing (p=0.0039, β=0.0014, SE=0.0005), and getting up (p=0.0066, β=0.0014, SE=0.0005) (Table S5). Lastly, smoking duration was used as continuous variable, which showed a positive relationship for smoking with walking and balance (p=0.0003, β=0.0046, SE=0.0012), freezing (p=6×10^−5^, β=0.0048, SE=0.0012), and getting up (p<1×10^−5^, β=0.0061, SE=0.0012) (Table S5).

**Table 1.** Motor symptoms associated with environmental factors. Percentage of patients stratified by smoking status and coffee consumption and motor symptoms. P-value: Regression model to predict the respective motor symptoms adjusted for covariates by including AAE, gender and disease duration (time between AAO and current age) and for smoking lung disease as comorbidity in the model →glm(formula = MotorSymptomYes ∼ AAE + Gender + DiseaseDuration + EnvFactorYes (+Comorbidity), family = gaussian, data = data).

|  |  | Smoking |  |  | Coffee |  |  |
| --- | --- | --- | --- | --- | --- | --- | --- |
|  |  | yes | no | p-value | yes | no | p-value |
| Tremor | yes | 79.1%<br>(n=1672) | 77.4%<br>(n=2585) | 0.1537 | 78.1%<br>(n=3082) | 79.0%<br>(n=886) | 0.5631 |
|  | no | 20.9%<br>(n=441) | 22.6%<br>(n=756) |  | 21.9%<br>(n=864) | 21.0%<br>(n=236) |  |
| Speech | yes | 71.5%<br>(n=1248) | 69.4%<br>(n=1925) | 0.2074 | 69.4%<br>(n=2261) | 70.4%<br>(n=670) | 0.1520 |
|  | no | 28.5%<br>(n=866) | 30.6%<br>(n=1418) |  | 30.6%<br>(n=1687) | 29.6%<br>(n=453) |  |
| Saliva and Drooling | yes | 69.5%<br>(n=1132) | 68.9%<br>(n=1655) | **0.0165** | 68.2%<br>(n=1971) | 71.6%<br>(n=587) | 0.0797 |
|  | no | 30.5%<br>(n=982) | 31.1%<br>(n=1688) |  | 31.8%<br>(n=1977) | 28.4%<br>(n=536) |  |
| Chewing and Swallowing | yes | 30.4%<br>(n=804) | 28.9%<br>(n=1133) | **0.0004** | 27.9%<br>(n=1361) | 31.2%<br>(n=433) | **0.0454** |
|  | no | 69.6%<br>(n=1310) | 71.1%<br>(n=2210) |  | 72.1%<br>(n=2587) | 68.8%<br>(n=690) |  |
| Walking and Balance | yes | 59.0%<br>(n=1469) | 57.6%<br>(n=2303) | 0.3995 | 57.3%<br>(n=2690) | 59.7%<br>(n=803) | 0.1114 |
|  | no | 41.0%<br>(n=644) | 42.4%<br>(n=1038) |  | 42.7%<br>(n=1256) | 40.3%<br>(n=319) |  |
| Freezing | yes | 53.5%<br>(n=643) | 49.5%<br>(n=964) | **0.0272** | 49.9%<br>(n=1099) | 52.3%<br>(n=350) | 0.2046 |
|  | no | 46.5%<br>(n=1470) | 50.5%<br>(n=2377) |  | 50.1%<br>(n=2847) | 47.7%<br>(n=772) |  |
| Getting up | yes | 38.0%<br>(n=1511) | 33.9%<br>(n=2317) | 0.1397 | 34.5%<br>(n=2740) | 38.6%<br>(n=790) | 0.4579 |
|  | no | 62.0%<br>(n=602) | 66.1%<br>(n=1024) |  | 65.5%<br>(n=1206) | 61.4%<br>(n=332) |  |

In addition, smoking status showed an impact on non-motor symptoms, especially on symptoms related to mood (Table S6 and Table S7). When modeled in a logistic regression to predict non-motor symptoms (Supplementary text), with covariates smoking (binary)/duration/dosage, AAE, gender, and disease duration (time between AAO and current age), smoking exhibited a positive relationship with unexplained pains (p<1×10^−5^, β=0.0624, SE=0.0135), problems remembering (p=0.0001, β=0.0540, SE=0.0141), feeling sad (p<1×10^−5^, β=0.0794, SE=0.0140), anxiety (p<1×10^−5^, β=0.0667, SE=0.0133), changed interest in sex (p=0.0013, β=0.0424, SE=0.0132), and light-headedness (p=0.0005, β=0.0490, SE=0.0140), when smoking was used as a binary yes-no indication (Table S8).

When cigarettes smoked per day were used as continuous variable for which non-smokers were set to zero, smoking showed a positive relationship with unexplained pains (p=0.0056, β=0.0015, SE=0.0005), problems remembering (p=0.0024, β=0.0017, SE=0.0005), feeling sad (p=0.0007, β=0.0018, SE=0.0005), anxiety (p=0.0001, β=0.0020, SE=0.0005), and light-headedness (p=0.0090, β=0.0014, SE=0.0005) (Table S8).

Lastly, the smoking duration was used as continuous variable. Smoking duration presented a positive relationship with unexplained pains (p=0.0131, β=0.0033, SE=0.0013), problems remembering (p=0.0016, β=0.0043, SE=0.0014), feeling sad (p=0.0444, β=0.0027, SE=0.0013), and changed interest in sex (p=0.0376, β=0.0026, SE=0.0013) (Table S8).

### Caffeine

Patients with iPD who drank coffee regularly had a later AAO (n=3993; median AAO=61.9 years; IQR=54.7-67.6) compared to patients with iPD who did not drink coffee at all (n=1133; median AAO=59.4 years; IQR=52.1-65.6) (p<0.0001) (Fig. 2A and Table S3). Investigation of a possible coffee dosage effect revealed that the number of cups of coffee per week was associated with AAO, although the effect size was small (n=4028, r=0.10, p<0.0001) (Fig. 2B). Longer coffee drinking duration also showed a positive correlation with AAO (n=2051, r=0.69, p<0.0001) (Fig. 2C). We investigated whether AAE contributed to the correlation between coffee drinking and AAO. When modeled in a linear regression to predict AAO, AAE (p<1×10^−5^, β>0.8239, SE<0.0122), coffee drinking dosage (p=8×10^−5^, β=0.0309, SE=0.0078), and coffee drinking duration (p<1×10^−5^, β=0.1268, SE=0.0083) remained in the model as independent predictors.

**Fig. 2.**
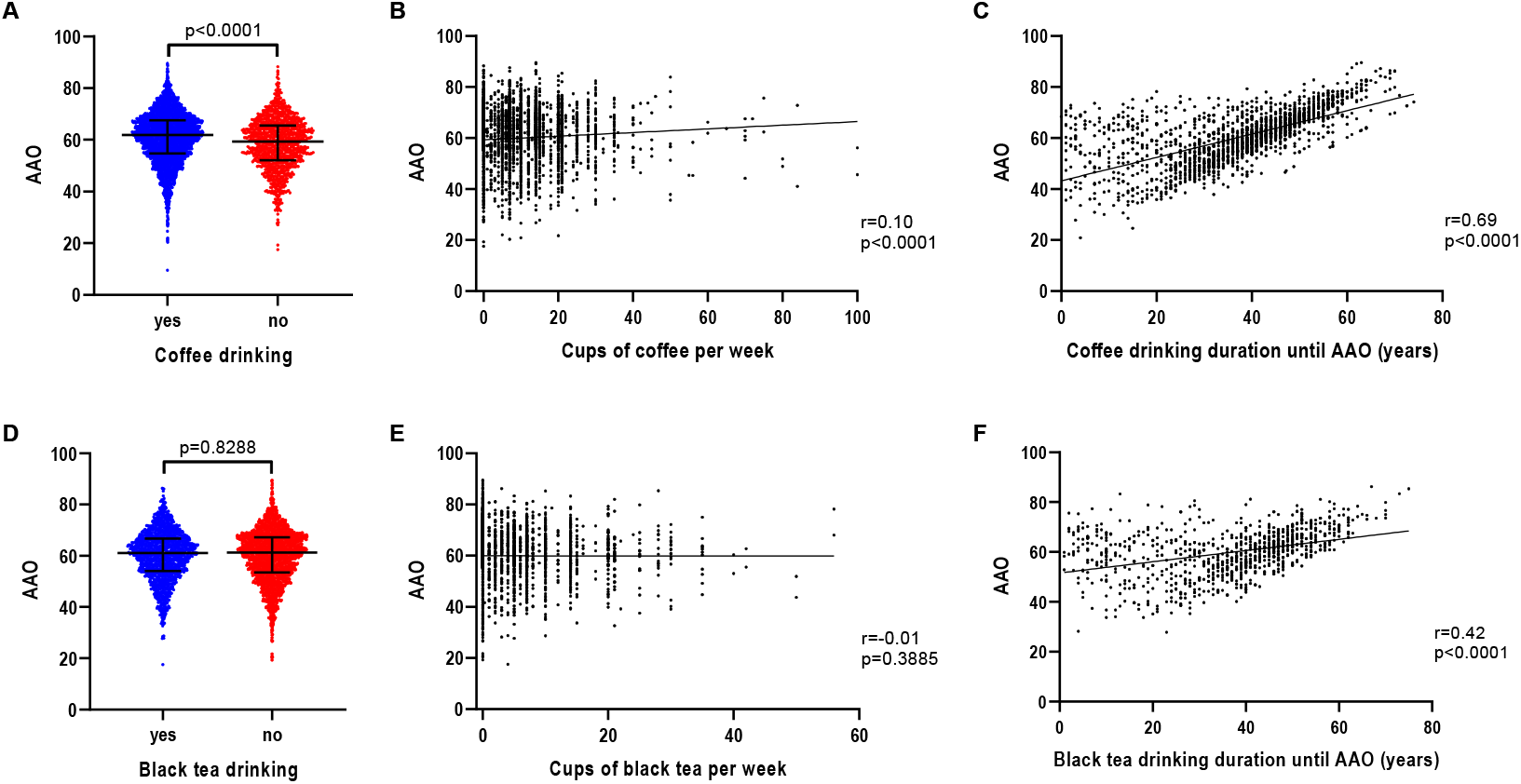
Association of AAO and caffeine consumption, caffeine drinking intensity and caffeine drinking duration in iPD. **(a)** Scatter plot of AAO of patients with iPD stratified by coffee consumption. Median values and interquartile ranges (IQR) are depicted. **(b)** Correlation between number of cups of coffee per week and AAO of patients with iPD. **(c)** Correlation between number of years of coffee drinking until AAO and AAO of patients with iPD. **(d)** Scatter plot of AAO of patients with iPD stratified by black tea consumption. **(e)** Correlation between number of cups of black tea per week and AAO of patients with iPD. **(f)** Correlation between number of years of black tea drinking until AAO and AAO of patients with iPD. P-value: Exploratory Mann Whitney U-test was performed for pairwise comparisons; non-parametric Spearman correlation and simple linear regression analyses were used to assess interactions between variables; p = Spearman’s exploratory p-value, r= Spearman’s rank correlation coefficient

We performed a sensitivity analysis to evaluate more potential predictors of AAO. When modeled in a linear regression to predict AAO (Supplementary text), with covariates coffee drinking duration/dosage, AAE, gender, and disease duration (time between AAO and current age), a positive relationship with coffee drinking duration (p=0.0006, β=0.0088, SE=0.0026) was revealed. In contrast, coffee drinking dosage did not predict AAO (p=0.8459, β=-0.0004, SE=0.0020). However, a positive relationship for AAO with AAE (p<1×10^−5^, β>0.9835, SE<0.0038) and a negative relationship with disease duration (p<1×10^−5^, β<-0.9376, SE<0.0064) were also observed.

We further investigated the impact of coffee drinking on self-reported motor and non-motor features (Table 1 and Table S6). When modeled in a logistic regression to predict the occurrence of motor symptoms (Supplementary text), with covariates coffee drinking (binary)/duration/dosage, AAE, gender, and disease duration (time between AAO and current age), coffee drinking demonstrated a negative relationship with chewing and swallowing (p=0.0454, β=-0.0327, SE=0.0163), when coffee drinking was used as a binary yes-no indication (Table S5). When the number of cups of coffee per week were used as continuous variable for which non-drinkers were set to zero, coffee drinking dosage showed a positive relationship with tremor (p=0.0435, β=0.0013, SE=0.0007) (Table S5). Lastly, the coffee drinking duration was used as continuous variable, but did not show an association with any of the motor symptoms (Table S5).

In contrast, coffee did not show an association with non-motor symptoms (p>0.05) (Table S6), when modeled in a logistic regression to predict non-motor symptoms (Supplementary text), with covariates coffee drinking (binary)/duration/dosage, AAE, gender, and disease duration (time between AAO and current age), when coffee drinking was used as a binary yes-no indication and also when the coffee drinking duration was used as continuous variable (Table S8). When the number of cups of coffee per week were used as continuous variable for which non-drinkers were set to zero, coffee drinking dosage showed a positive relationship with unexplained pains (p=0.0085, β=0.0020, SE=0.0008), feeling sad (p=0.0391, β=0.0017, SE=0.0008), changed interest in sex (p=0.0201, β=0.0018, SE=0.0008) and light-headedness (p=0.0325, β=0.0017, SE=0.0008) (Table S8).

In contrast to the findings for coffee and AAO, black tea drinking was not observed to be associated with AAO (Fig. 2D and Table S3). There was also no association between the number of cups of black tea per week and AAO (n=3781, r=-0.01, p=0.3885) (Fig. 2E). However, there was a positive correlation of black tea drinking duration with AAO (n=930, r=0.42, p<0.0001) (Fig. 2F).

### Aspirin

When investigating the effect of anti-inflammatory medication on AAO of patients with iPD, aspirin showed the greatest difference in AAO. Patients with iPD, who reported the use of aspirin, had a five-year later AAO (n=1003; median AAO=64.0 years; IQR=57.9-69.0) compared to patients who did not take aspirin (n=1989; median AAO=59.1 years; IQR=51.8-64.9) (p<0.0001) (Fig. 3A and Table S3). The difference in AAO for ibuprofen-based non-aspirin medication was small (ibuprofen users: n=1087; median AAO=60.6 years; IQR=53.2-66.3; ibuprofen non-users: n=2008; median AAO=61.1 years; IQR=54.2-67.0; p=0.0345) or in the case of other anti-inflammatory medication we found no association at all (other anti-inflammatory drug users: n=498; median AAO=61.5 years; IQR=54.0-66.9; other anti-inflammatory drug non-users: n=2393; median AAO=60.7 years; IQR=53.7-66.6; p=0.2495). The number of aspirin pills per week was associated with AAO (n=2849, r=0.23, p<0.0001) (Fig. 3B). Likewise, the aspirin intake duration showed an association with AAO (n=577, r=0.23, p<0.0001) (Fig. 3C), indicating a later AAO the longer the patients took aspirin before disease onset. When examining the effect of AAE on aspirin intake and AAO by modeling in a linear regression to predict AAO, both AAE (p<1×10^−5^, β>0.9195, SE<0.0199) and aspirin intake duration (p=0.0171, β=0.0319, SE=0.0133) remained in the model but the aspirin dosage diminished as independent predictor (p=0.0974, β=0.0315, SE=0.0190).

**Fig. 3.**
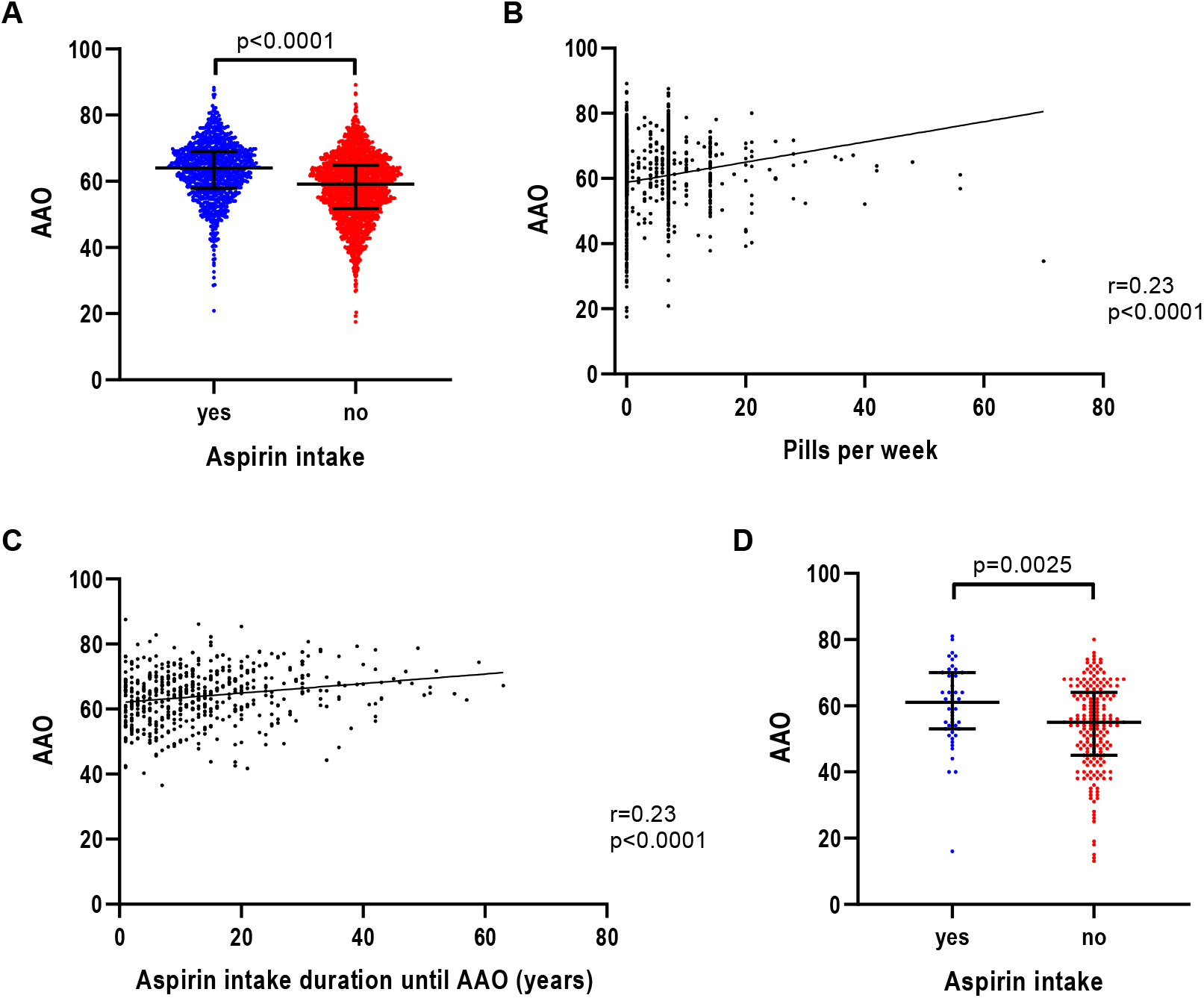
Association of AAO and aspirin intake, aspirin intake intensity and aspirin intake duration in iPD. **(a)** Scatter plot of AAO of patients with iPD stratified by aspirin intake. Median values and interquartile ranges (IQR) are depicted. **(b)** Correlation between number of aspirin pills per week and AAO of patients with iPD. **(c)** Correlation between number of years of aspirin intake until AAO and AAO of patients with iPD. **(d)** Scatter plot of AAO of patients with iPD from the EPIPARK replication cohort stratified by aspirin intake. P-value: Exploratory Mann Whitney U-test was performed for pairwise comparisons; non-parametric Spearman correlation and simple linear regression analyses were used to assess interactions between variables; p = Spearman’s exploratory p-value, r= Spearman’s rank correlation coefficient

To evaluate more potential predictors of AAO we performed a sensitivity analysis. When modeled in a linear regression to predict AAO (Supplementary text), with covariates aspirin intake duration/dosage, AAE, gender, disease duration (time between AAO and current age) and potential comorbidities (heart diseases/arthritis/back pain/surgeries with anesthesia), the aspirin intake duration was not associated with AAO (p>0.8399, β>-0.0009, SE<0.0044). In addition, dosage was also not associated with AAO (p>0.4236, β<0.0038, SE<0.0048). However, in the models that included the aspirin intake duration as covariate, a positive relationship for AAO with AAE (p<1×10^−5^, β>0.9850, SE=0.0066) and arthritis (p=0.0444, β=0.1973, SE=0.0979) were observed, while disease duration (p<1×10^−5^, β<-0.9198, SE<0.0128), as well as heart diseases (p=0.0378, β=-0.2185, SE=0.1049) presented a negative relationship with AAO. When including the aspirin intake dosage in the model instead of the duration, a positive relationship for AAO with AAE (p<1×10^−5^, β>0.9928, SE<0.0027), and a negative relationship with disease duration (p<1×10^−5^, β<-0.9551, SE=0.0046), heart diseases (p=0.0500, β=-0.1346, SE=0.0687) and surgeries with anesthesia (p=0.0024, β=-0.2384, SE=0.0786) were observed. Given that the effect of aspirin diminished as an independent predictor after adjusting for covariates, we did not further investigate the impact of aspirin use on motor and non-motor features.

### Replication cohort

Since the aspirin and PD AAO association, although not dependable in the regression models, has not been investigated and published previously, we utilized a separate German iPD cohort to investigate further. In the EPIPARK cohort, patients with iPD who reported the use of at least one aspirin pill per week over a minimal period of one month had a more than six year later AAO (n=49; median AAO=61.0 years; IQR=53.0-70.0) compared to patients who did not take aspirin (n=188; median AAO=55.0 years; IQR=45.0-64.0) (p=0.0025) (Fig. 3D).

To examine the effect of aspirin and AAE on AAO we used a linear regression model to predict AAO, showing that AAE (p<7×10^−5^, β>0.9942, SE<0.1400) remained in the model but again the effect of aspirin duration (p=0.1792, β=0.3375, SE=0.2261) as well as of aspirin dosage (p=0.0660, β=0.4440, SE=0.2401) diminished as independent predictor.

## Discussion

We found an association between the general intake of aspirin, intake duration and number of pills per week with later AAO. We further replicated our findings concerning aspirin in a separate German iPD cohort (EPIPARK) [26]. However, in a linear regression model to predict AAO, aspirin diminished as independent predictor, raising the question that aspirin may possibly be confounded. Testing several potential comorbidities that could be connected to aspirin use revealed an association between heart diseases, arthritis and surgeries with anesthesia and AAO. In fact, PD patients with heart diseases had a later AAO compared to patients with no heart diseases. The same applies to patients with arthritis. Moreover, the percentage of aspirin users was higher in PD patients with heart diseases or arthritis than in PD patients without these diseases. The effect on PD AAO was not extended to other NSAIDs in the Fox Insight cohort. Nevertheless, we identified an association between heart diseases and surgeries with anesthesia and AAO, when performing a regression model including AAE, gender, disease duration, ibuprofen intake dosage and other comorbidities. The clinical effect of NSAIDs is still subject to controversial discussion. While some studies indicate a protective effect of NSAIDs or at least an association with PD [20, 27], others may see a neuroprotective potential of NSAIDs but not an association with PD at the population level [28, 29]. No other studies have explored aspirin and AAO in a large iPD cohort so far (Fig. S1). In addition to the novel findings, we replicated previous associations for smoking and caffeine with AAO in PD, summarized in a systematic literature review (Fig. S1 and Table S1) [7, 8, 10-13, 15-18, 23, 24]. This effect was further supported by multivariate regression models. However, when evaluating the independence of smoking and coffee drinking from AAE by pairwise correlations, it showed that smoking dosage, coffee drinking dosage, and coffee drinking duration remained in the model as independent predictors, but smoking duration did not. The opposite directionality for the effect of smoking duration on AAO that was seen in the sensitivity analysis can also be explained by the correlation between smoking duration and AAE that destabilizes the model. Moreover, the effect of smoking and coffee drinking dosage on AAO diminished when including more covariates, which is likely due to small effect sizes. In our exploratory analysis of the Fox Insight cohort, more patients with iPD felt depressed, anxious, helpless, worthless or hopeless when smoking or former smoking compared to non-smokers. There is evidence of an established relationship between smoking and mental health [30, 31]. Still, the causal effect remains unclear. In other words, smoking itself may promote depression and anxiety, or patients with depression are just more likely to smoke and have greater difficulty quitting [32, 33]. Moreover, the underlying cause of more severe motor symptoms in smokers remains unclear. Since smokers in this cohort were on average older than non-smokers, but had a shorter disease duration, we would rather have expected less severe motor symptoms. To examine whether smoking has an actual impact on motor and non-motor symptoms, we will perform longitudinal studies in the future to determine a possible long-lasting effect of tobacco use and smoking on PD-related symptoms and will further separate former smokers with PD and current smokers with PD to predict this long-lasting effect. Furthermore, the mechanism of action needs to be investigated in functional studies. This might explain the discrepancy between later AAO but more severe motor and non-motor symptoms in smokers.

Although the effect size for the number of cups of coffee was relatively low, the coffee drinking duration showed a strong effect, consistent with previous studies [27, 34-36], which was also verified in the regression models. Consistent with our study, six other studies showed that coffee drinkers have a later PD onset, one study showed an opposite effect (Table S1). Caffeine is the speculated reason for the protective effect of coffee. In the Fox Insight cohort, coffee drinkers showed less severe motor symptoms compared to patients who did not drink coffee, that are in line with previous studies on tremor and dyskinesia [37]. However, a positive relationship with tremor was observed when the number of cups of coffee per week was used as continuous variable. The opposite direction of trends can be explained by different sample sizes, because not all coffee drinkers provided data on their drinking dosage. Additionally, the effect size was very small. There was a lack of association between non-motor symptoms and coffee consumption in general in our study, however we found a positive relationship between non-motor symptoms and the coffee drinking dosage. Nevertheless, the effect of coffee consumption and potential long-term effects need to be investigated in further longitudinal studies. Long-term effects of coffee drinking may positively impact memory, cognition and verbal retrieval [16, 38, 39]. Black tea had a more modest association with AAO, likely due to a lower amount of caffeine.

One strength of our study was the large sample size that provided sufficient power to assess lifestyle factors and PD onset, but also allows small magnitude associations to show significance. In addition, online self-report data collections offer many possibilities to promote epidemiological research because of convenience and accessibility for the participants and researchers. A previous study compared self-reported demographic characteristics, symptoms, medical history, and PD medication use of the Fox Insight PD cohort to other in-person observational research study cohorts [40]. They found that patterns of responses to patient-reported assessments that were obtained online on the PD cohort of the Fox Insight study were similar to PD cohorts assessed in-person. Patient-reported outcomes are becoming increasingly important to research, therapeutic development and healthcare delivery, which was already investigated in another previous study on Fox Insight [41]. However, due to the self-report assessments data may also contain more subjective perceptions that are difficult to standardize. Nevertheless, these findings may help to acquire a better understanding of this complex disease that can be used to developed specific therapeutic strategies.

This study is a comprehensive assessment of smoking, caffeine and aspirin intake on the onset of iPD, as well as clinical symptoms. Besides replicating previous findings in a large self-report American cohort, novel associations of comorbidities linked to aspirin use with PD AAO were observed. These findings are so far only exploratory, however, they set the stage for future longitudinal assessments on these factors and PD clinical features.

## Data Availability

Data used in the preparation of this article were obtained from the Fox Insight database (https://foxden.michaeljfox.org/insight/explore/fox.jsp) on 18/10/2020. For up-to-date information on the study, visit https://foxden.michaeljfox.org/insight/explore/fox.jsp.

## Acknowledgments

The Fox Insight Study (FI) is funded by The Michael J. Fox Foundation for Parkinson’s Research. We would like to thank the Parkinson’s community for participating in this study to make this research possible. This study was funded by The Michael J. Fox Foundation for Parkinson’s Research (Analysis of Patient-Reported Outcomes from Fox Insight). JT, MK, and CK are supported by the DFG (FOR 2488).

