## Supplementary Data for "Coffee, smoking and aspirin are associated with age at onset and clinical severity in idiopathic Parkinson’s disease"

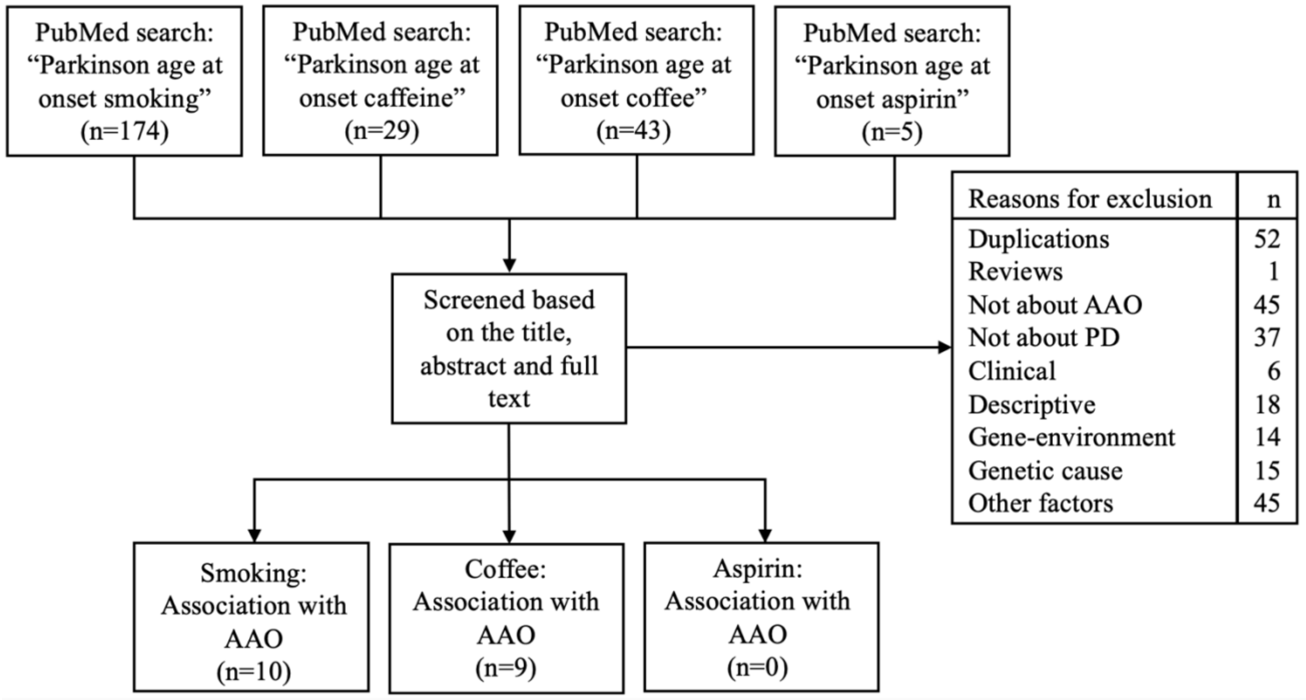


**Fig. S1** Work flow of the literature search in PubMed. We searched for literature via PubMed that was published before April 14, 2021. We used the free text search terms "Parkinson age at onset smoking", resulting in 174 articles, "Parkinson age at onset caffeine", resulting in 29 articles, "Parkinson age at onset coffee", resulting in 43 articles and "Parkinson age at onset aspirin", resulting in 5 articles. These were screened based on the title, abstract and full text, excluding all articles not directly investigating smoking, coffee drinking or aspirin intake and their influence on AAO in PD. Reasons for exclusion and the number of excluded articles are shown. In the end, 10 articles were obtained that described a relationship between AAO and smoking, and 9 articles describing a relationship between AAO and coffee, with an overlap of 6 articles. There was no study that investigated the association between AAO and aspirin. Duplications: duplicates between the different search terms; Not about AAO: articles investigating smoking, caffeine/coffee or aspirin, but not AAO; Not about PD: articles about other diseases or about symptoms or parts of PD; Clinical: studies on clinical applications, diagnosis or therapy of PD; Descriptive: articles describing a population, symptoms, features or comorbidities of PD; Gene-environment: studies investigating gene-environment interactions and the influence of environmetal factors on specific genotypes; Genetic cause: only investigating a genetic factor influencing PD, no lifestyle factors; Other factors: studies investigating other environmental factors than smoking, coffee/caffeine or aspirin in PD

**Table S1** Publications on AAO and smoking/coffee of the literature search in PubMed

| Authors, Year (PMID) | n (PD/control) | n (smokers with PD/ non-smokers with PD) | n (coffee drinkers with PD/ coffee non-drinkers with PD) | Effect on AAO |
|---|---|---|---|---|
| Yahalom et al., 2020 (32310186) [1] | 225/0<br><br>(65 with LRRK2 G2019S PD and 60 with GBA N370S PD) | 98 /127<br><br>(26 with LRRK2 G2019S PD and 39 mutation carriers and 27 with GBA N370S PD and 33 mutation carriers) | 199 /25<br><br>(56 with LRRK2 G2019S PD and 9 mutation carriers and 53 with GBA N370S PD and 6 mutation carriers) | **Smoking:**<br>- Smoking associated with AAO (p = 0.032)<br>→ Later AAO |
| | | | | **Coffee:**<br>- Consumption level of coffee (p=0.001) significantly associated with PD AAO<br>→ Later AAO the higher the amount of coffee |
| Benedetti et al., 2000 (11087780) [2] | 196/196 | NA | NA | **Smoking:**<br>- Ever-smokers: median AAO 70 years<br>- Never-smokers: median AAO 71.5 years<br>→ Similar AAO |
| | | | | **Coffee:**<br>- Coffee-drinkers: median AAO 72 years<br>- Non-drinkers median AAO 64 years<br>→ Later AAO |
| Wijeyekoon et al., 2017 (29057010) [3] | 144/102 | NA | NA | **Smoking:**<br>- Ever-smoking in males associated with delayed AAO (p=0.048)<br>→ Later AAO (in males) |
| | | | | **Coffee:**<br>- Regular coffee drinking associated with later AAO (p<0.001)<br>→ Later AAO |

| | | | | |
|---|---|---|---|---|
| Kandinov et al., 2009 (18434232) [4] | 278/0 | 111/167 | 180/98 | **Smoking:**<br>- Smokers: 1-9 pack-years: mean AAO 57.1 years, ≥10 pack-years: mean AAO 61.2 years<br>- Non-smokers: mean AAO 57.2 years<br>- A higher amount of cigarettes smoked per day, showed a later AAO<br>→ Later AAO (when ≥10 pack-years) |
| | | | | **Coffee:**<br>- Coffee-drinkers: <2 daily cups: mean AAO 58 years, 2-3 daily cups: mean AAO 57.6 years, >3 daily cups: mean AAO 55 years<br>- Coffee non-drinkers: mean AAO 59.5 years<br>→ Earlier AAO (with dosage effect) |
| Lüth et al., 2020 (32875616) [5] | 342/57<br><br>(142 with LRRK2 G2019S PD) and 57 mutation carriers) | 112 /199<br><br>(41 smokers with LRRK2 G2019S PD and 85 non-smokers with LRRK2 G2019S PD) | 182 /130<br><br>(62 coffee drinkers with LRRK2 G2019S PD and 63 coffee non-drinkers with LRRK2 G2019S PD) | **Smoking:**<br>- LRRK2 PD:<br>Smokers: median AAO 60 years<br>Non-smokers: median AAO 52 years ($p=0.0215$)<br>- iPD:<br>Smokers: median AAO 55 years,<br>Non-smokers: median 53.5 years ($p=0.7906$)<br>- Number of cigarettes per day correlated with AAO ($p=0.0296$) as well as smoking duration ($p<0.0001$)<br>→ Later AAO (in LRRK2 PD) |
| | | | | **Coffee:**<br>- LRRK2 PD:<br>Coffee-drinkers: median AAO 55 years<br>Non-drinkers: median AAO 52 years ($p=0.5439$)<br>- iPD:<br>Coffee-drinkers: median AAO 55 years<br>Non-drinkers: median AAO 52 years ($p=0.3279$)<br>→ Trend to a later AAO, but no significant difference |

| | | | | |
|---|---|---|---|---|
| Maher et al., 2002 (11781409) [6] | 396/0 | NA | NA | **Smoking:**<br>- Among siblings who smoked, pack-years of smoking was related to later age at onset (p=0.0001)<br>→ Later AAO |
| | | | | **Coffee:**<br>- The mean age at onset did not differ according to exposure to coffee<br>→ Similar AAO |
| Haack et al., 1981 (7304554) [7] | 237/474 | 87/150 | 223/14<br><br>(coffee or tea) | **Smoking:**<br>- Smokers: mean AAO 52.7 years (men: 54.4 years, women: 42.5)<br>- Non-smokers: mean AAO 57.8 years (men: 58.9 years, women: 55.4)<br>→ Earlier AAO |
| Kuopio et al., 1999 (10584666) [8] | 123/246 | NA | NA | **Smoking:**<br>- Ever-smokers: AAO 65.0 years<br>- Never-smokers: AAO 61.8 years (p=0.051)<br>→ Later AAO |
| De Reuck et al., 2005 (15792818) [9] | 512/0 | 184/328 | NA | **Smoking:**<br>- Ever-smokers: mean AAO 65.9 years<br>- Never-smokers: mean AAO 62.4 years (p=0.001)<br>→ Later AAO |
| Gigante et al., 2017 (28988683) [10] | 262/0 | 111/151 | NA | **Smoking:**<br>- Ever-smokers: mean AAO 61.69 years<br>- Never-smokers: mean AAO 59.28 years (p=0.03)<br>→ Later AAO |
| Gigante et al., 2018 (29362953) [11] | 83/0 | 12/71 | 71/12 | **Coffee:**<br>- Number of coffee drinking years associated with a significant increase in AAO (p<0.001)<br>→ Later AAO (dosage effect) |

| | | | | |
|---|---|---|---|---|
| Cho et al., 2018 (29449185) [12] | 196/0 | NA | 136/60 | **Coffee:**<br>- Coffee drinkers: mean AAO 63.4 years<br>- Non-coffee drinkers: mean AAO 67.3 years ($p=0.008$)<br>→ Later AAO |
| Tan et al. 2007 (18075470) [13] | 418/468 | 81/337 | 401/17 | **Coffee:**<br>- Significant association between caffeine intake and the onset of PD ($p=2.01x10^{-5}$)<br>- Dosage effect, showing a later AAO the higher the caffeine consumption<br>→ Later AAO (with dosage effect) |

**Table S2** Demographics of the Fox Insight participants

| Full Cohort (n=35,963) | Patients with PD |
|---|---|
| **Male (%)** | 18,349 (51.0%) |
| **Female (%)** | 14,528 (40.4%) |
| **Ethnicity:** | |
| **White/Caucasian (%)** | 32,332 (89.9%) |
| **Black/African American (%)** | 369 (1.0%) |
| **American Indian/Alaska Native (%)** | 393 (1.1%) |
| **Asian (%)** | 691 (1.9%) |
| **Native Hawaiian/Other Pacific Islander (%)** | 47 (0.1%) |
| **Hispanic/Latino/Spanish Origin (%)** | 1,692 (4.7%) |
| **Mean AAO (SD)** | 60.4 (11.0) |
| **Mean AAE (SD)** | 65.7 (10.2) |
| **Median AAO (IQR)** | 61.3 (53.6-68.1) |
| **Median AAE (IQR)** | 66.7 (59.6-72.6) |

**Supplementary text:**

***Fox Insight study:***

The Fox Insight study is an ongoing online, longitudinal health study of people with and without PD with targeted enrollment set to at least 125,000 individuals [14]. The data is a rich data set facilitating discovery, validation, and reproducibility in PD research. The dataset is generated through routine longitudinal assessments (health and medical questionnaires evaluated at regular cycles); one-time health and disease questionnaires about symptoms, daily activities, and other factors; and, in a subgroup of people with PD, genetic data collection. Qualified researchers can explore, analyze, and download patient-reported outcomes (PROs) data and PD-related genetic variants at https://foxden.michaeljfox.org. The full Fox Insight genetic data set, including approximately 650,000 single nucleotide polymorphisms (SNPs) per participant, can be requested separately with institutional review.

Fox Insight participants were 18 years of age or older and provided informed consent. In the process of registration, participants were divided into two groups, PD patients and controls, the latter were asked about new diagnoses every three months. PD patients responded to health, non-motor assessments, motor assessments, quality of life, and lifestyle questionnaires. These questionnaires based on the Movement Disorders Society – Unified Parkinson's disease Rating Scale (MDS-UPDRS) Part II, the Non-Motor Symptoms Questionnaire (NMSQ), and the Geriatric Depression Scale (GDS). The PD-RFQ-U on "Smoking and Tobacco" questionnaire was used to evaluate smoking, the PD-RFQ-U on "Caffeine" to evaluate coffee drinking and black tea drinking, and the PD-RFQ-U on "Anti-inflammatory Medication History" for anti-inflammatory drug intake. The surveys on "Your Movement Experiences" (MDS-UPDRS Part II; The scores range from 1 to 5, with higher scores indication more severe symptoms) and "Your Non-Movement Experiences" (NMSQ; Scores of 0 and 1) were used to assess the association between smoking, coffee and black-tea drinking, and aspirin intake with motor and non-motor symptoms. Finally, the surveys on "Your Current Health" and "Your Mood" (GDS; Scores of 0 and 1) were used to examine the association between smoking and mood, anxiety and depression. All of these data were self-reported by the patients.

For each environmental or lifestyle factor the corresponding datasets were downloaded from the FoxDEN website (https://foxden.michaeljfox.org/insight/explore/fox.jsp) (log:18/10/2020).

***Statistical analysis:***

For a first statistical analysis, non-parametric Mann-Whitney U test was performed to compare the distribution of AAO between different groups. For correlation analyses, non-parametric Spearman correlations and linear regression analyses were used to assess correlations and interactions between variables (GraphPad Software Inc., San Diego, CA, USA). For a more in-depth analysis, we performed multilinear regression models to investigate the relationship between environmental factors, age, disease duration, motor/non-motor symptoms and potential comorbidities (IBM SPSS Statistics).

*Regression model investigating AAO, AAE, environmental factors (duration/dosage):*

To evaluate whether the environmental factors were correlated to age, this multiple regression model used the AAO as dependent variable and AAE and duration or dosage (for which non-users were set to zero) of each environmental factor as covariates.

→ glm(formula = AAO ~ AAE + EnvFactorDosage, family = gaussian, data = data)
→ glm(formula = AAO ~ AAE + EnvFactorDuration, family = gaussian, data = data)

*Regression model investigating AAO, AAE, gender, disease duration, environmental factors (duration/dosage) and comorbidities:*

To evaluate potential confounders, this multiple regression model adjusted for more covariates, using the AAO as dependent variable and AAE, gender, disease duration (time between AAO and current age) and duration or dosage (for which non-users were set to zero) of each environmental factor as covariates. For the investigation of smoking and aspirin, several potential comorbidities (lung diseases; heart diseases, arthritis, back pain and surgeries with anesthesia) were investigated (IBM SPSS Statistics).

→ glm(formula = AAO ~ AAE + Gender + DiseaseDuration + EnvFactorDosage (+Comorbidity), family = gaussian, data = data)
→ glm(formula = AAO ~ AAE + Gender + DiseaseDuration + EnvFactorDuration (+Comorbidity), family = gaussian, data = data)

*Regression model investigating environmental factors and motor/non-motor symptoms adjusted for AAE, gender, disease duration:*
To assess the relationship between environmental factors and motor/non-motor symptoms, multiple regression models were estimated to predict the respective motor/non-motor symptoms. To examine potential confounders, we adjusted for covariates by including AAE, gender and disease duration (time between AAO and current age) and for smoking lung disease as comorbidity in the model. The regarding environmental factor was handled in three different ways: In a first set of analyses, it was used binary as yes-no indication for the environmental factor, in the second set of analyses, the dosage was used as continuous variable, for which non-users were set to zero and in the third set of analyses, the duration was used as continuous variable (IBM SPSS Statistics). For the investigation of motor symptoms, motor symptoms were dichotomized into yes (scores greater than 1) versus no (scores equal to 1), since continuous variables could not be used since they were not normally distributed.
→ glm(formula = MotorSymptomYes ~ AAE + Gender + DiseaseDuration + EnvFactorYes (+Comorbidity), family = gaussian, data = data)
→ glm(formula = MotorSymptomYes ~ AAE + Gender + DiseaseDuration + EnvFactorDosage (+Comorbidity), family = gaussian, data = data)
→ glm(formula = MotorSymptomYes ~ AAE + Gender + DiseaseDuration + EnvFactorDuration (+Comorbidity), family = gaussian, data = data)
→ glm(formula = NonMotorSymptomYes ~ AAE + Gender + DiseaseDuration + EnvFactorYes, family = gaussian, data = data)
→ glm(formula = NonMotorSymptomYes ~ AAE + Gender + DiseaseDuration + EnvFactorDosage, family = gaussian, data = data)
→ glm(formula = NonMotorSymptomYes ~ AAE + Gender + DiseaseDuration + EnvFactorDuration, family = gaussian, data = data)

**Table S3** Association of environmental factors and AAO. Median AAO stratified by tobacco use, coffee consumption, black tea consumption and aspirin intake

| | yes | no | p-value |
|---|---|---|---|
| | | **Tobacco** | |
| n | 2148 | 3375 | NA |
| Median AAO (IQR) | 63.5 (56.1-69.1) | 60.8 (53.7-66.7) | <0.0001 |
| | | **Coffee** | |
| n | 3993 | 1133 | NA |
| Median AAO (IQR) | 61.9 (54.7-67.6) | 59.4 (52.1-65.6) | <0.0001 |
| | | **Black tea** | |
| n | 1719 | 2449 | NA |
| Median AAO (IQR) | 61.0 (54.1-66.7) | 61.3 (53.5-67.2) | 0.8228 |
| | | **Aspirin** | |
| n | 1003 | 1989 | NA |
| Median AAO (IQR) | 64.0 (57.9-69.0) | 59.1 (51.8-64.9) | <0.0001 |

**Table S4** Cumulative number of patients. The motor symptom scores range from 1 to 5 (1: normal (no problems), 2: slight, 3: mild, 4: moderate, 5: severe)

| | | n | | | |
|---|---|---|---|---|---|
| | Score | **Smokers** | **Non-smokers** | **Coffee drinkers** | **Coffee non-drinkers** |
| Tremor | 1 | 441 | 756 | 864 | 236 |
| | 2 | 932 | 1454 | 1751 | 471 |
| | 3 | 550 | 853 | 1017 | 295 |
| | 4 | 166 | 240 | 274 | 103 |
| | 5 | 24 | 38 | 40 | 17 |
| Speech | 1 | 866 | 1418 | 1687 | 453 |
| | 2 | 498 | 759 | 919 | 246 |
| | 3 | 515 | 821 | 954 | 298 |
| | 4 | 211 | 315 | 357 | 108 |
| | 5 | 24 | 30 | 31 | 18 |
| Saliva and Drooling | 1 | 982 | 1688 | 1977 | 536 |
| | 2 | 323 | 493 | 612 | 167 |
| | 3 | 434 | 681 | 797 | 236 |
| | 4 | 306 | 406 | 458 | 159 |
| | 5 | 69 | 75 | 104 | 25 |
| Chewing and Swallowing | 1 | 1310 | 2210 | 2587 | 690 |
| | 2 | 622 | 932 | 1104 | 340 |
| | 3 | 60 | 79 | 91 | 36 |
| | 4 | 120 | 117 | 161 | 57 |
| | 5 | 2 | 5 | 5 | 0 |
| Walking and Balance | 1 | 644 | 1038 | 1256 | 319 |
| | 2 | 890 | 1491 | 1738 | 499 |
| | 3 | 264 | 366 | 441 | 135 |
| | 4 | 264 | 383 | 434 | 141 |
| | 5 | 51 | 63 | 77 | 28 |
| Freezing | 1 | 1470 | 2377 | 2847 | 772 |
| | 2 | 348 | 543 | 623 | 179 |
| | 3 | 146 | 219 | 251 | 80 |
| | 4 | 108 | 147 | 169 | 65 |
| | 5 | 41 | 55 | 56 | 26 |
| Getting up | 1 | 602 | 1024 | 1206 | 332 |
| | 2 | 960 | 1546 | 1843 | 484 |
| | 3 | 358 | 524 | 604 | 204 |
| | 4 | 150 | 202 | 237 | 81 |
| | 5 | 43 | 45 | 56 | 21 |

**Table S5** Motor symptoms associated with environmental factors in regression models. P-value: Multivariate regression to predict the respective motor symptoms adjusted for covariates by including AAE, gender and disease duration (time between AAO and current age) and for smoking lung disease as comorbidity in the model

→ glm(formula = MotorSymptomYes ~ AAE + Gender + DiseaseDuration + EnvFactorYes (+Comorbidity), family = gaussian, data = data)

→ glm(formula = MotorSymptomYes ~ AAE + Gender + DiseaseDuration + EnvFactorDosage (+Comorbidity), family = gaussian, data = data)

→ glm(formula = MotorSymptomYes ~ AAE + Gender + DiseaseDuration + EnvFactorDuration (+Comorbidity), family = gaussian, data = data)

| | **Smoking** | | | **Coffee** | | |
|---|---|---|---|---|---|---|
| | p-value | | | p-value | | |
| | Yes/No | Dosage | Duration | Yes/No | Dosage | Duration |
| Tremor | 0.1537 | 0.8110 | 0.4699 | 0.5631 | **0.0435** | 0.3862 |
| Speech | 0.2074 | 0.1779 | 0.0596 | 0.1520 | 0.7770 | 0.2328 |
| Saliva and Drooling | **0.0165** | 0.0507 | 0.1621 | 0.0797 | 0.8448 | 0.9677 |
| Chewing and Swallowing | **0.0004** | **0.0004** | 0.1248 | **0.0454** | 0.8460 | 0.4846 |
| Walking and Balance | 0.3995 | **0.0320** | **0.0003** | 0.1114 | 0.8335 | 0.5607 |
| Freezing | **0.0272** | **0.0039** | **$6x10^{-5}$** | 0.2046 | 0.6042 | 0.9856 |
| Getting up | 0.1397 | **0.0066** | **$<1x10^{-5}$** | 0.4579 | 0.5285 | 0.6278 |

**Table S6** Non-motor symptoms associated with environmental factors. Percentage of patients stratified by smoking status and coffee consumption and non-motor symptoms. P-value: Multivariate regression to predict the respective non-motor symptoms adjusted for covariates by including AAE, gender and disease duration (time between AAO and current age)
→ glm(formula = NonMotorSymptomYes ~ AAE + Gender + DiseaseDuration + EnvFactorYes, family = gaussian, data = data)

| | | **Smoking** | | | **Coffee** | | |
|---|---|---|---|---|---|---|---|
| | | yes | no | p-value | yes | no | p-value |
| Constipation | yes | 55.1% (n=1174) | 53.6% (n=1801) | 0.2036 | 53.5% (n=2127) | 57.0% (n=643) | 0.0955 |
| | no | 44.9% (n=957) | 46.4% (n=1559) | | 46.5% (n=1847) | 43.0% (n=486) | |
| Unexplained Pains | yes | 40.0% (n=852) | 35.7% (n=1199) | **$<1x10^{-5}$** | 37.2% (n=1480) | 37.5% (n=423) | 0.1608 |
| | no | 60.0% (n=1278) | 64.3% (n=2161) | | 62.8% (n=2494) | 62.5% (n=706) | |
| Problems Remembering | yes | 50.6% (n=1078) | 44.9% (n=1510) | **0.0001** | 47.0% (n=1869) | 46.8% (n=528) | 0.9866 |
| | no | 49.4% (n=1052) | 55.1% (n=1850) | | 53.0% (n=2105) | 53.2% (n=601) | |
| Feeling Sad | yes | 53.9% (n=1148) | 48.0% (n=1611) | **$<1x10^{-5}$** | 50.2% (n=1993) | 50.3% (n=567) | 0.2533 |
| | no | 46.1% (n=982) | 52.0% (n=1746) | | 49.8% (n=1980) | 49.7% (n=560) | |
| Anxiety | yes | 38.7% (n=825) | 34.2% (n=1149) | **$<1x10^{-5}$** | 36.2% (n=1438) | 35.9% (n=405) | 0.2181 |
| | no | 61.3% (n=1305) | 65.8% (n=2208) | | 63.8% (n=2535) | 64.1% (n=722) | |
| Changed Interest in Sex | yes | 35.7% (n=761) | 32.4% (n=1089) | **0.0013** | 34.6% (n=1376) | 32.3% (n=364) | 0.1007 |
| | no | 64.3% (n=1369) | 67.6% (n=2268) | | 65.4% (n=2597) | 67.7% (n=763) | |
| Light-headedness | yes | 45.7% (n=974) | 41.4% (n=1390) | **0.0005** | 43.4% (n=1723) | 41.7% (n=470) | 0.2805 |
| | no | 54.3% (n=1156) | 58.6% (n=1966) | | 56.6% (n=2250) | 58.3% (n=656) | |

**Table S7** Mood associated with smoking status. Percentage of patients stratified by smoking status and symptoms related to mood. P-value: Multivariate regression to predict the respective mood symptoms adjusted for covariates by including AAE, gender and disease duration (time between AAO and current age)

→ glm(formula = MoodSymptomYes ~ AAE + Gender + DiseaseDuration + SmokingYes, family = gaussian, data = data)

| | | **Smoking** | | |
|---|---|---|---|---|
| | | yes | no | p-value |
| Depression | yes | 29.8% (n=627) | 24.2% (n=799) | **$<1x10^{-5}$** |
| | no | 70.2% (n=1475) | 75.8% (n=2509) | |
| Anxiety | yes | 31.5% (n=661) | 27.2% (n=900) | **$1x10^{-5}$** |
| | no | 68.5% (n=1440) | 72.8% (n=2409) | |
| Dropped many activities and interests | yes | 40.1% (n=837) | 33.0% (n=1087) | **$<1x10^{-5}$** |
| | no | 59.9% (n=1252) | 67.0% (n=2204) | |
| Life feels empty | yes | 16.0% (n=333) | 13.2% (n=433) | **0.0004** |
| | no | 84.0% (n=1751) | 86.8% (n=2858) | |
| Getting bored often | yes | 29.4% (n=615) | 22.3% (n=734) | **$<1x10^{-5}$** |
| | no | 70.6% (n=1475) | 77.7% (n=2558) | |
| Being afraid something bad could happen | yes | 25.0% (n=521) | 22.5% (n=739) | **0.0022** |
| | no | 75.0% (n=1559) | 77.5% (n=2551) | |
| Feeling helpless often | yes | 21.8% (n=453) | 17.8% (n=584) | **$2x10^{-5}$** |
| | no | 78.2% (n=1628) | 82.2% (n=2703) | |
| Prefer staying at home | yes | 54.6% (n=1136) | 50.0% (n=1644) | **0.0007** |
| | no | 45.4% (n=946) | 50.0% (n=1645) | |
| Feeling to have more memory problems than other people | yes | 28.4% (n=592) | 25.0% (n=823) | **0.0019** |
| | no | 71.6% (n=1494) | 75.0% (n=2472) | |
| Feeling pretty worthless | yes | 17.0% (n=354) | 14.1% (n=462) | **0.0002** |
| | no | 83.0% (n=1727) | 85.9% (n=2821) | |
| Feeling that situation is hopeless | yes | 14.8% (n=308) | 11.3% (n=372) | **$<1x10^{-5}$** |
| | no | 85.2% (n=1767) | 88.7% (n=2912) | |

**Table S8** Non-motor symptoms associated with environmental factors in regression models. P-value: Multivariate regression to predict the respective motor symptoms adjusted for covariates by including AAE, gender and disease duration (time between AAO and current age) in the model
→ glm(formula = NonMotorSymptomYes ~ AAE + Gender + DiseaseDuration + EnvFactorYes, family = gaussian, data = data)
→ glm(formula = NonMotorSymptomYes ~ AAE + Gender + DiseaseDuration + EnvFactorDosage, family = gaussian, data = data)
→ glm(formula = NonMotorSymptomYes ~ AAE + Gender + DiseaseDuration + EnvFactorDuration, family = gaussian, data = data)

| | **Smoking** | | | **Coffee** | | |
|---|---|---|---|---|---|---|
| | p-value | | | p-value | | |
| | Yes/no | Dosage | Duration | Yes/no | Dosage | Duration |
| Constipation | 0.2036 | 0.6288 | 0.6234 | 0.0955 | 0.9160 | 0.4410 |
| Unexplained Pains | **$<1x10^{-5}$** | **0.0056** | **0.0131** | 0.1608 | **0.0085** | 0.0890 |
| Problems Remembering | **0.0001** | **0.0024** | **0.0016** | 0.9866 | 0.5652 | 0.5355 |
| Feeling Sad | **$<1x10^{-5}$** | **0.0007** | **0.0444** | 0.2533 | **0.0391** | 0.5334 |
| Anxiety | **$<1x10^{-5}$** | **0.0001** | 0.3919 | 0.2181 | 0.1881 | 0.5979 |
| Changed Interest in Sex | **0.0013** | 0.2010 | **0.0376** | 0.1007 | **0.0201** | 0.6895 |
| Light-headedness | **0.0005** | **0.0090** | 0.1328 | 0.2805 | **0.0325** | 0.1948 |
